# The untapped health and climate potential of cycling in France: a national assessment from individual travel data

**DOI:** 10.1101/2023.04.04.23288155

**Authors:** Emilie Schwarz, Marion Leroutier, Audrey De Nazelle, Philippe Quirion, Kévin Jean

## Abstract

**Background:** Promoting active modes of transportation such as cycling may generate important public health, economic, and climate mitigation benefits. We aim to assess mortality and morbidity impacts of cycling in a country with relatively low levels of cycling, France, along with associated monetary benefits; we further assess the potential additional benefits of shifting a portion of short trips from cars to bikes, including projected greenhouse gas emissions savings.

**Methods:** Using individual data from a nationally-representative mobility survey, we described the French 2019 cycling levels by age and sex. We conducted a burden of disease analysis to assess the incidence of five chronic diseases (breast cancer, colon cancer, cardio-vascular diseases, dementia, and type-2 diabetes) and numbers of deaths prevented by cycling, based on national incidence and mortality data and dose-response relationships from meta-analyses. We assessed the corresponding direct medical cost savings and the intangible costs prevented based on the value of a statistical life year. Lastly, based on individual simulations, we assessed the likely additional benefits of shifting 25% of short (<5km) car trips were shifted to cycling.

**Findings:** The French adult (20-89 years) population was estimated to cycle on average of 1min 17sec pers^-1^.day^-1^ in 2019, with important heterogeneity across gender and age. This yielded benefits of 1,919 (uncertainty interval, UI: 1,101-2,736) premature deaths and 5,963 (UI: 3,178-8,749) chronic disease cases prevented, with males enjoying nearly 75% of these benefits. Direct medical costs prevented were estimated at €191 million (UI: 98-285) annually, while the corresponding intangible costs were nearly 25 times higher (€4.8 billion, UI: 3.0-6.5). We estimated that in average, €1.02 (UI: 0.59-1.62) of intangible costs were prevented for every km cycled. Shifting 25% of short car trips to biking would yield approximatively a 2-fold increase in death prevented, while also generating important CO_2_ emission reductions (0.257 MtCO2e, UI: 0.231-0.288).

**Interpretation:** In a country of low- to moderate cycling culture, cycling already generates important public health and health-related economic benefits. Further development of active transportation would increase these benefits while also contributing to climate change mitigation targets.

**Funding:** This research received no specific grant from any funding agency in the public, commercial, or not-for-profit sectors.

## Introduction

Insufficient physical activity is responsible for a substantial global burden, with an approximatively 7% of all-cause deaths attributable. This burden is even more pronounced in Europe, with an approximate 10% of all-cause deaths and 30% of direct health-care costs of non-communicable diseases and mental health conditions attributable to physical inactivity ^1,2^. In high-income Western countries, the prevalence of insufficient physical activity reaches more than 40% while showing no decreasing trends over time ^3^. While the disquieting situation results in part from individual behaviors and choices, collective and societal decisions bear an essential responsibility, especially with regards to transport-related physical activity ^4^. Active travel offers a unique opportunity to boost physical activity by encouraging seamless integration in daily activities, requiring little to no additional time commitments and costs to the individual ^5^. Despite clear benefits of the promotion of utilitarian cycling, most countries do not use it to its full potential: in temperate regions such as the UK or France, the modal share of cycling remains below 3%, far from the 15 to 27% rates obtained in countries such as the Netherlands or Denmark ^6,7^.

Collective choices regarding mobility also largely contribute to climate change, which is widely recognized as one the biggest public health issues of the upcoming decades ^8^. In Europe, the transportation sector represents the second largest contributor of greenhouse gas emissions ^9^, and the first in France where it accounts for 31% of the national emissions, among which approximately 50% are attributable to cars ^10^. Therefore, promoting modal shift from cars to active transportation, such as cycling, represents a relevant response to both the public health burden of physical inactivity and the urgent need to cut down emissions.

While the public health benefits of cycling have already been assessed in a large variety of contexts ^11,12^, national assessments based on representative mobility data are less common ^13^. Moreover, few burden of disease studies have assessed its impact in terms of both mortality and morbidity or have estimated the associated direct medical and intangible costs avoided ^14,15^. Such assessments can contribute meaningfully to intense debates currently ongoing about the future of mobility in a context of ambitious decarbonisation targets and energy crises in many high-income Western countries such as France.

Our aim in this study was to demonstrate how cycling can contribute to health promotion in a Western country with relatively low cycling rates. We first assessed the benefits associated with current cycling levels in France in terms of prevented mortality and morbidity, based on nationally-representative individual mobility data from 2019. Secondly, we assessed the benefits that could be associated with shifting a portion of short trips currently made by car to bike trips.

## Methods

### Mobility data

Individual-level mobility data were accessed from the 2019 *Enquête mobilité des personnes* (“People’s mobility survey”) ^16^. This survey is conducted every 10 years and the most recent data represents the mobility of the metropolitan French population in 2019, before the onset of the Covid-19 pandemic. 13,825 individuals aged 5 years and older were asked face to face about their travels the day before. Distances travelled are consolidated using geographic information systems, based on reported departure and arrival location. The sampling design and sampling weights ensure that the survey is representative of travel behaviours across weekdays and weekends in France. We analyzed all cycling trips, regardless of travel purpose and week day. Distances cycled were converted into exposure time (i.e. minutes of cycling per day) considering an average cycling speed of 14.9 km.h^-1^, independently of gender and age, and similarly to recent studies including two conducted in France ^17–19^. As e-bikes were not frequently used in 2019, for simplicity, we assumed the same speed and metabolic equivalent of task (MET) values for e-bikes than for classical bikes.

### Morbi-mortality and demographic data

While insufficient physical activity has been linked to a range of health outcomes ^20–26,^ 5 diseases were selected for this health impact assessment (HIA) based on data availability regarding: i) an established dose-response relationship with physical activity estimated in a meta-analysis, ii) national incidence, iii) estimates of corresponding medical costs based on health coverage data. The diseases considered were: breast cancer in females, colon cancer, cardiovascular disease, dementia, and type 2 diabetes (Table 1). We also assumed a protective effect of cycling on all-cause mortality as documented by the meta-analysis by Kelly et al ^27^. As this meta-analysis considered all-cause mortality, potential increases in exposure to air pollution or injury are implicitly controlled. We thereafter use the term “morbi-mortality” to refer to these five chronic diseases and all-cause mortality. For each morbi-mortality event, the relative risk (RR) identified in the literature was scaled for a reference volume of 11.25 MET.hours (supplementary Table 1).

**Table 1:** Data sources, medical costs, disability weights and dose-response functions used in the analysis to assess the health and health-related economic impact of physical activity.

| Morbi-mortality event | Incidence data | Related Medical costs (per case)* | Disability weight | RR, for 100 min cycling, ie 11.25 MET.hours** (95% CI) | RR Reference |
| --- | --- | --- | --- | --- | --- |
| Breast Cancer (females) | National Cancer Institute | 46,968 € | 0.068 | 0.90 (0.87-0.95) | Monninkhof et al., 2007 |
| Colon Cancer | National Cancer Institute | 26,716 € | 0.093 | Males: 0.93 (0.88-0.99)<br>Females: 0.94 (0.91-0.99) | Harris et al., 2009 |
| Cardiovascular Disease | Long-term condition coverage | 20,938 € | 0.052 | 0.92 (0.90-0.95) | Hamer & Chida, 2008 |
| Dementia | Long-term condition coverage | 22,748 € | 0.152 | 0.90 (0.86-0.95) | Hamer & Chida, 2009 |
| Type 2 Diabetes | Long-term condition coverage | 36,514 € | 0.068 | 0.81 (0.72-0.89) | Jeon et al., 2007 |
| All-cause mortality | INSEE | - | 1 | 0.90 (0.87,.94) | Kelly et al., 2014 |

RR: Relative risk, CI: confidence interval.
\* Disease-specific medical costs are estimated based on the average duration of disease <sup>31</sup>.
\*\* 11.25 MET.hours corresponds to the international recommendations of at least 150 minutes of moderate-intensity aerobic physical activity per week.
See Supplementary Table 1 for the initial values of RR, the references used for physical activity and the scaling for RR for 100min cycling.

France-specific incident cases for cardiovascular disease, dementia, and type 2 diabetes were determined for the year 2018 from a database of new beneficiaries of long-term illness coverage (*Affectations Longues Durées*, ALD) by the French National health insurance system, using the most recent data available. The 2019 incidence data for breast cancer and colon cancer were taken from the French National Cancer Institute. Deaths, death rates, life-expectancy and population count estimates for 2019 were taken from the National Institute of Statistics and Economic Studie ^28^ (Table 1). All the morbi-mortality data mentioned here were accessed and used as age- and sex-specific data.

### Health impact modelling

The health benefits of 2019 levels of cycling were assessed compared to a counterfactual “zero cycling” scenario in which trips did not entail any physical activity.

For each individual surveyed, time exposure to cycling (if any) was converted into a percentage reduction in the age-specific risk for each morbi-mortality event. Therefore, for each event, we estimated the reduced individual risk attributable to cycling as compared to the zero cycling counterfactual scenario. Following the Health Economic Assessment Tool (HEAT) approach, we assumed linear dose-response functions (DRF) for the mortality and morbidity effects of cycling while disregarding the levels of other types of physical activity, and we capped the all-cause mortality reduction at 45% ^29^. The age range considered for reductions in morbidity and mortality was 20-89 years. We disregarded younger and older ages because of the scarcity of evidence for the health effects of physical activity in these age ranges for these specific outcomes. For each morbi-mortality event, by summing up the reduction in individual probability of event across the study sample, we estimated the total number of events that have been averted by cycling. Survey weights were applied in the calculation in order to obtain nationally representative figures across weekdays and weekends.

### Burden of Disease

Based on the estimated age-specific numbers of morbi-mortality events prevented, we estimated the corresponding Disability-Adjusted Life Years (DALY) prevented by adding up the years of life lost (YLL) and the years lived with disability (YLD). Disability weights as those provided by the Global Burden of Disease (GBD) are generally specific to the level of severity of the disease considered. In order to obtain disease-specific average disability weights which accounts for the severity spectrum of the disease, we divided disease-specific YLD-estimates by disease-specific prevalence estimates obtained from the 2019 GBD^30^. For the DALY estimation, disability weights were applied from the disease onset over the expected remaining years to live.

### Health economic evaluation

We evaluated the monetarized benefits of 2019 French cycling levels by considering two different definitions of costs. We first estimated the *medical costs* associated with chronic diseases included in the analyses. Disease-specific medical costs were estimated based on expenses reimbursed by all health insurance schemes (available at: https://data.ameli.fr/explore/dataset/depenses/information/). These expenses, expressed in Euros 2018 (Table 1), include outpatient care, hospitalization in public or private healthcare facilities, and daily allowances. Total disease-specific medical costs were estimated from annual costs assuming an average duration of disease, therefore the calculation does not consider age at disease onset ^31^. For each disease, the average duration was estimated under the assumption of stationary prevalence, by the ratio between the total number of prevalent cases and the total number of incident cases, provided by the 2019 GBD (Supplementary Table 2). Such evaluation of the tangible medical costs prevented by cycling does not include the benefits on mortality. Therefore, we also estimated the intangible costs prevented by cycling using the standard value of statistical life year (VSLY) that is recommended in France for the socioeconomic evaluation of public investments ^32^. Estimates of intangible costs prevented by cycling thus rely on the VSLY of 133k€, expressed in Euros 2019.

### Uncertainty analysis

Central values and 95% confidence intervals for cycling exposure were estimated while accounting for the survey sampling design and weights using the R package *survey* ^33^.

Uncertainty surrounding the RR relating cycling and morbi-mortality events were characterized by a log-normal distribution constructed for each outcome based on the RR central values and the lower and upper bounds of the 95% confidence interval provided in the literature. We then combined the uncertainties using a Monte Carlo approach where we independently sampled a RR value for each morbi-mortality event. We randomly sampled 100 combinations of RR values. Based on a given set of randomly-sampled RR values, the mean value and the standard-error for each outcome were estimated and results were combined across sets of RR combinations using the Rubin’s rule ^34^. The distributions corresponding to each of the 100 replications were then combined together to generate a posterior distribution for each outcome, from which we computed the 2.5% and 97.5% percentiles to obtain 95% uncertainty intervals (UI).

### Modal shift scenario

Lastly, based on individual-level simulations, we assessed the potential additional public health benefits associated with a modal shift scenario in which a share of short car trips (defined as <5km) would be shifted to cycling. To do so, we randomly selected 25% of respondents whose car trips were all <5km, while also reporting no cycling trips, and estimated the corresponding exposure time to cycling had these trips been cycled. We then re-conducted the steps of the HIA described above to assess the health and health-related economic benefits associated, while assuming no substitution effect of cycling on other types of physical activity. We performed 100 random draws of individuals whose short car trips were re-allocated to bike trips. For each of these random selections, we computed the DALY prevented together with the 95% UI following the steps previously described. Results of each replication were combined to generate a posterior distribution for each outcome and compute 95% UI. Lastly, we estimated the CO_2_ emissions averted in this modal shift scenario, assuming that the 2019 French car fleet emitted on average 124 gCO_2_.km^-1^ ^35^. In a sensitivity analysis, we reproduced the same steps while using 10km, instead of 5km, as a cut-off to define short car trips, 25% of which would be shifted to cycling.

## Results

In 2019, among the French population of 20-89 year-olds (population size, 49.0 million), the cumulative kilometers biked in France added up to 4.64 (95% CI: 3.28-6.00) billion kilometers, of which 6.23% were cycled on e-bikes. This represented a per-capita average distance biked of 0.32 (95% CI: 0.27-0.37) km.pers^-1^.day^-1^, corresponding to an average exposure time of 1min 17sec pers^-1^.day^-1^. The proportion of the population reporting any cycling trip on a given day, accounting for differences in weekends and weekdays, was 3.12% (95% CI: 2.64-3.61%), with variations according to age and sex (Figure 1). In all age groups, the proportion of cyclist was higher in males vs. females, and the average distance cycled among cyclists was higher in male vs. female. This resulted in 72.2% (95% CI: 60.7%-82.6%) of all cycling distances being biked by males.

**Figure 1:**
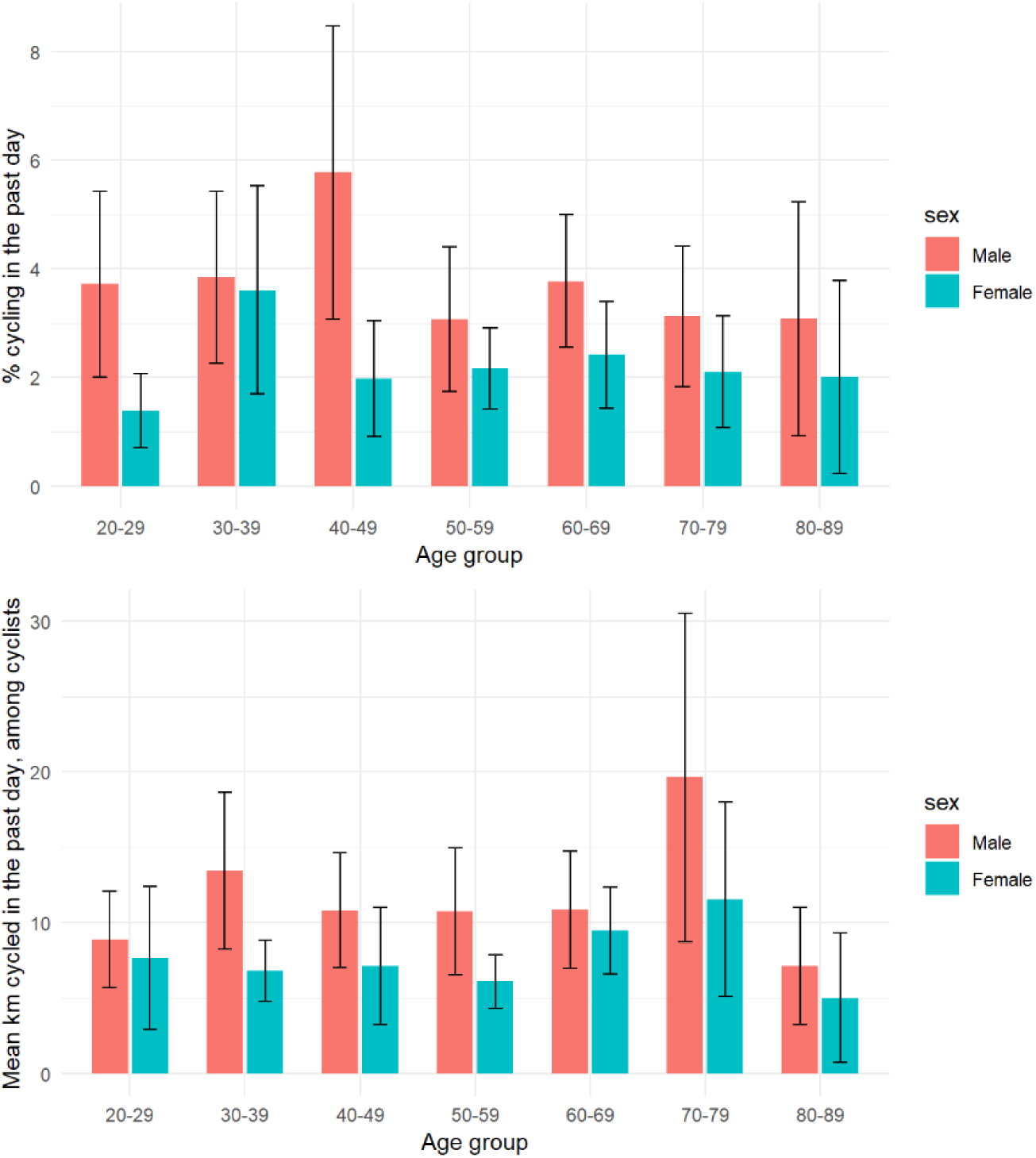
Proportion of the French adult population reporting any bike trip any cycle trip a day, accounting for differences in weekends and weekdays (top), and mean distance cycled (km) in the past day among those reporting any bike trip (bottom) according to sex and age. *Enquête mobilité des personnes,* France, 2019. Black lines represent 95% confidence intervals of the gender- and age-specific percentage (top) and mean (bottom).

### Health impact assessment

Based on the observed cycling levels, we estimated that 1,919 (UI: 1,101-2,736) premature deaths and 5,963 (UI: 3,178-8,749) chronic diseases were prevented in 2019 in France due the protective effect of biking physical activity. The chronic disease with the greatest number of cases prevented was type-2 diabetes (n=3,743, UI: 1,576-5,912), 3743.838 followed by CVD (n=1,578, UI: 778-2,378). Of all morbi-mortality events prevented, 74.9% benefited males (Figure 2). On average, cycling reduced the yearly mortality for the total adult population by 0.68% (UI: 0.47% - 0.87%).

**Figure 2:**
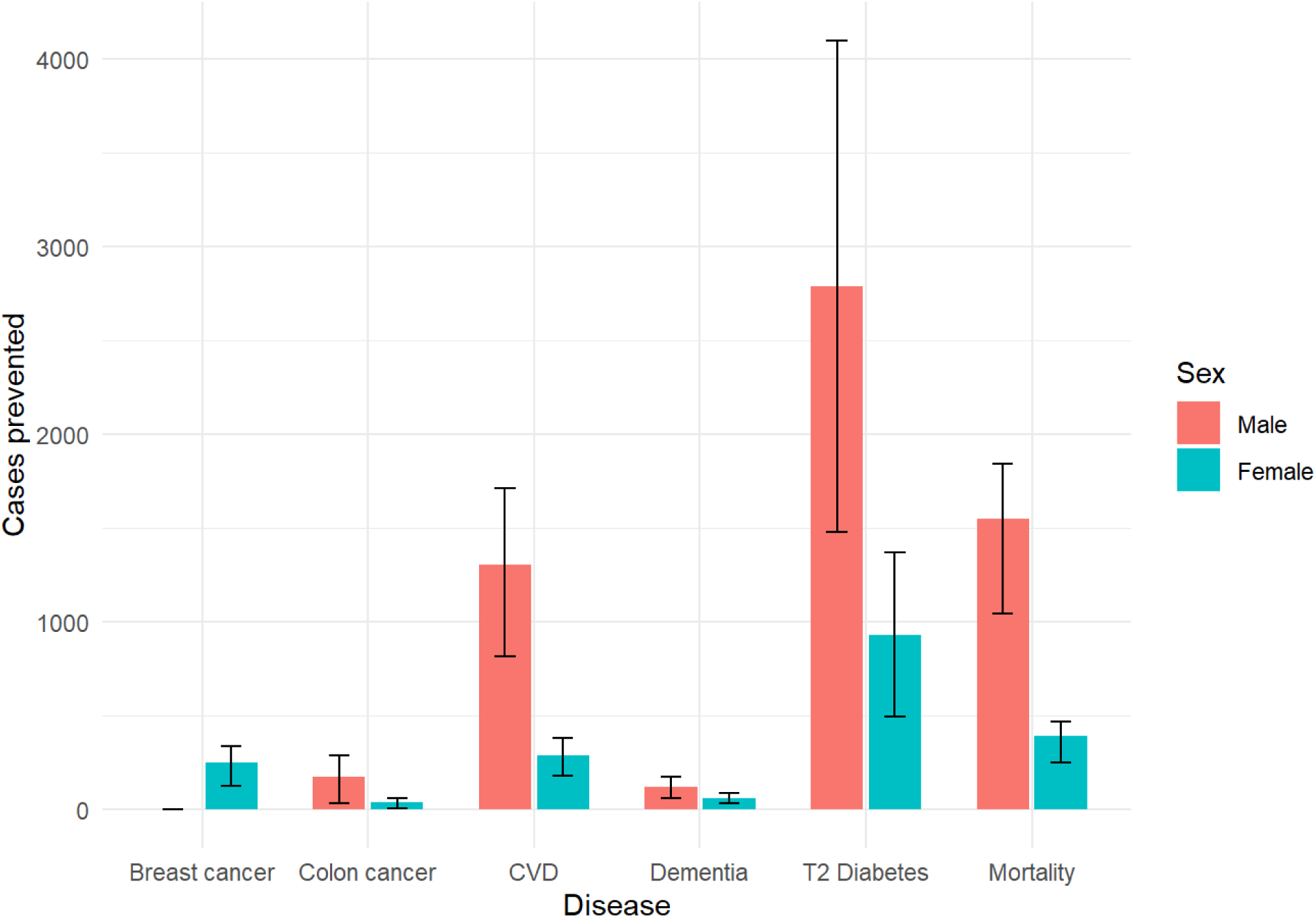
Chronic diseases and mortality prevented by the physical activity due to cycling in France among adults aged 20-89 years, 2019. Black lines represent uncertainty intervals.

We further estimated that overall, 35,135 DALYs (UI: 22,693 – 48,791) were prevented through cycling-related physical activity, of which 28,416 (80.9%) were driven by mortality (YLL) and 6,719 (19.1%) by morbidity (YLD) (Figure 3). Based on the average values of annual medical costs associated with each chronic disease considered here, we estimated that the 2019 levels of cycling saved €191 (UI: 98-285) million annual medical (tangible) costs. Based on the value of a statistical life year, combining intangible costs of mortality and morbidity, we estimated that cycling prevented €4.75 (UI: 3.02-6.49) billion, roughly 25 times the annual medical (tangible) costs. When related to the total kilometers traveled, this led to the estimate that each kilometer traveled by bike avoids an average of €1.02 (UI: 0.59-1.62) of intangible costs. Table 2 summarizes the burden and costs prevented for each morbi-mortality event.

**Figure 3:**
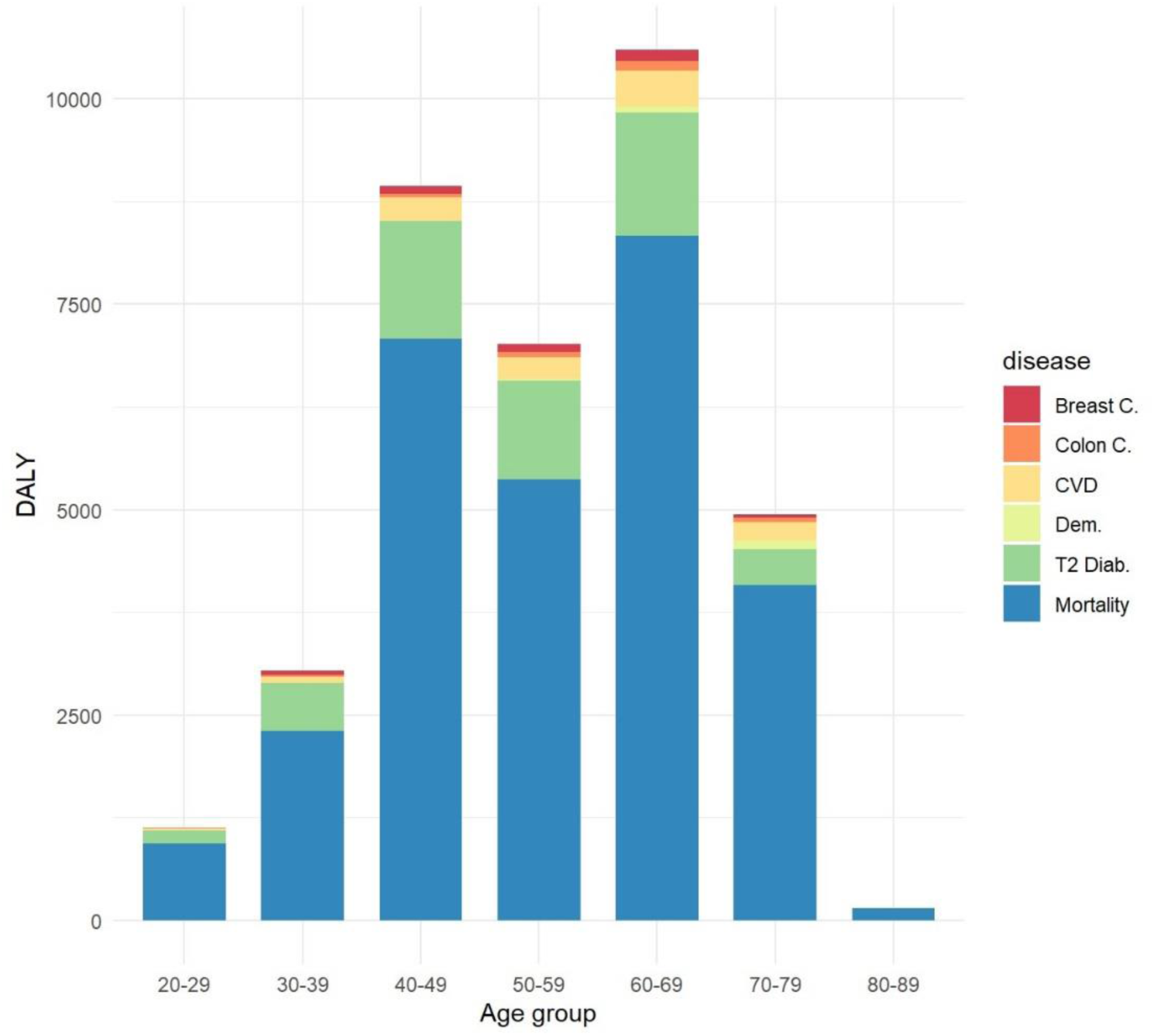
Disability-adjusted life years (DALY) prevented by physical activity due to cycling in France, 2019, according to age group. DALY associated with chronic diseases correspond to years of life with disability (YLD) and DALY associated with mortality correspond to years of life lost (YLL, in blue).

**Table 2:** Burden, tangible costs and intangible costs prevented by the physical activity due to cycling in France, 2019, for several morbi-mortality event, among adults aged 20-89 years.

| **Morbi-mortality event** | **Cases prevented in 2019 (Uncertainty interval, UI)** | **Medical (tangible) costs prevented in 2019, M€ (UI)** | **Intangible costs prevented in 2019, M€ (UI)** |
| --- | --- | --- | --- |
| Breast cancer | 254 (107-402) | 11.9 (5.0-18.9) | 58.2 (34.1-82.3) |
| Colon cancer | 205 (51-359) | 5.5 (1.4-9.6) | 35.5 (14.3-56.6) |
| Cardiovascular disease | 1578 (778-2378) | 33.0 (16.3-49.8) | 175.8 (113.6-238.0) |
| Dementia | 182 (68-295) | 4.1 (1.6-6.76) | 29.1 (15.9-42.2) |
| Diabetes Type 2 | 3,744 (1,576-5,912) | 136.76 (57.56-215.9) | 723.3 (382.1-1064.6) |
| Mortality | 1,919 (1,101-2,736) | Not Applicable | 3731.8 (2808.2-4655.4) |

Intangible costs are estimated based on a value of a statistical life year (VSLY) of 133k€, expressed in Euros 2019.

### Modal shift scenarios

The proportion of survey participants whose short car trips were all <5km car trip on a given day was 23.4%; 95% CI: 22.4-24.5) overall and was quite homogeneously distributed across age and sex, as was the mean length of these short car trips (2.42 km, 95% CI: 2.36-2.49) (Supplementary Figure 1).

Shifting 25% of short car trip to cycling would generate an additional 2.073 billion km cycled (UI: 1.864-2.314), ie an approximate 45% higher cycling exposure compared to the 2019 baseline. Due to the age distribution of drivers, this would translate into a approximate 2-fold deaths prevented compared to the 2019 cycling levels (1,822deaths prevented, UI: 1,010-2,633) (Table 3). This modal shift scenario would prevent €2.59 billion in terms of intangible costs (UI: 1.69-3.50), while also reducing CO2 emissions by 0. 257 mega-tons (UI: 0.231-0.288). Defining short trips on the basis of a distance twice as great (ie 10km instead of 5km, for short car trips, 25% of which could be shifted to cycling) resulted in health benefits which were approximatively two times greats (Supplementary Table 3).

**Table 3:** Climate, health and health-related economic benefits of cycling in France, 2019, and estimated impact of a modal shift scenario.

| **Outcome** | **Baseline estimates**<br>(Uncertainty interval, UI) | Incremental effect of shifting 25% of short (<5km) car trips to cycling (in addition to the baseline estimates) (UI) |
| --- | --- | --- |
| Yearly km cycled (billion) | 4.640<br>(3.284-5.996) | 2.073 (1.864-2.314) |
| CO<sub>2</sub> emissions prevented (Mto) | 0.575 (0.4070.743) <sup>1</sup> | 0.257 (0.231-0.288) |
| # of deaths prevented | 1,919 (1101-2736) | 1,822 (1,010-2,633) |
| # of chronic diseases prevented | 5,963 (3,178-8,749) | 3,410 (2,343-4476) |
| # DALYS prevented | 35,135 (22,693 – 48,791) | 19,493 (12,684-26,302) |
| Direct medical (tangible) costs savings (million €) | 191 (98-285) | 108 (71-144) |
| Intangible costs prevented (billion €) | 4.75 (3.02-6.49) | 2.59 (1.69-3.50) |
| Intangible costs prevented for every km cycled (€) | 1.02 (0.59-1.62) | 1.25 (0.82-1.73) |

Intangible costs are estimated based on the value of a statistical life year (VSLY).
<sup>1</sup> As compared to a counterfactual where individual would have done the same trips driving instead of cycling

## Discussion

In this study relying on a nationally-representative mobility survey, we show that, despite relatively low levels of cycling in France in 2019, physical activity due to cycling generated important public health benefits in terms of morbidity and mortality alleviated. These benefits were unequally distributed across sex, reflecting the cycling distribution, with males benefiting nearly 75% of the morbi-mortality events prevented. Of the 5 different chronic diseases considered, cycling levels mostly prevented cases of diabetes and CVD, and in all prevented €187 million in direct medical costs in 2019. Most of the public health benefits of cycling, however, resided in the mortality alleviated, which corresponded to approximately 80% of all DALY associated with cycling. When accounting for the VSLY, we estimate that the physical activity linked to the 2019 levels of cycling prevented nearly €5 billion of intangible costs annually. This corresponds to an approximate 1€ prevented for every km cycled. The prevented intangible costs associated with mortality largely outweighed the prevented direct medical costs linked to alleviated morbidity, at an approximate ratio of 25:1. Furthermore, we estimated that the annual number of death prevented by cycling could be doubled under a modest modal shift scenario of 25% short car trips to cycling, which would also lead to sizeable CO_2_ emissions reduction. To our knowledge, this study is the first to assess a feasible modal shift scenario based on simulations of individual-level data relying on a nationally-representative sample. These results confirm the relevance of promoting cycling both for planetary and public health.

Results from the 2019 wave of the French transport survey show that France remains within the countries with a low cycling culture ^36^. The French adult population reported cycling on average less than 10 minutes per week, which is low compared to the more than 70 minutes per week reported by the Dutch adult population ^13^. Despite repeated calls to decrease motorized trips to help tackle climate change and physical inactivity, cycling mode share for France remained at about 2.7% of all trips in France between 2008, the year of the previous wave of travel survey, and 2019. The bike modal remained the same (2.7%) between the two waves ^16^. This explains the relatively low levels of the health benefits of cycling in France as compared to other countries ^37^. In France, we estimate that cycling reduces the mortality risk by 0.6%, compared to 7.4% in the Netherlands ^13^. We showed that currently males are the main beneficiaries of cycling health benefits in France because they represent a large fraction of the cyclists. However, previous surveys have shown that females are equally represented in countries where cycling is normalized, providing an additional motivation for cycling promotion ^38^.

Our burden of disease approach allowed us to estimate the relative contribution of mortality and morbidity alleviated in the total health benefits of cycling. We found that cycling would prevent nearly 3 times more chronic diseases than deaths (approximatively 5,900 diseases and 1,800 deaths). This ratio was similar to the one reported in a previous HIA in Barcelona ^39,40^. However, the chronic conditions prevented are associated with relatively low disability weights (ranging between 0.05 and 0.15), which explains the large contribution of prevented mortality in the DALY. In terms of morbidity, we showed that the disease most frequently prevented were diabetes and cardio-vascular diseases. Previous studies reported that cycling also contributes to reducing mortality among adults with diabetes ^41^, making it a relevant intervention for both primary and secondary prevention. Due to the lack of reliable medical costs data, our estimate disregarded some diseases which were considered in some previous HIA of cycling, especially depression ^25,42^. Considering the high burden of depression in France (2019 national prevalence estimated at 3.46%, as compared to 4.65 for Type-2 Diabetes, for example^30^) and its considerable association with physical inactivity (Schuch et al estimated a risk significantly decreased by ∼30% for adults following international physical activity recommendations ^25^), this could have led to underestimate the health benefits of cycling we assessed here.

Our health economic evaluation contributes to current evidence on the substantial health and economic benefits of active transportation, even in countries of low- to moderate cycling culture^14,43^. Relying on nationally recommended monetarization values for the socio-economic evaluation of public investments, we show that current cycling practices already prevent nearly €5 billion in intangible costs annually. This represents an approximate €1 prevented for every kilometer cycled. This unit value is of a similar order of magnitude to those reported in other return on investment studies. For instance, in Queensland, Australia, it was reported to be AUD 0.67 (approximately €0.4) per kilometer ridden.^44^ Such substantial monetized health benefits associated with cycling contribute to building a strong case for significant investment in cycling.^45^

We further estimated the public and climate mitigation benefits of a modal shift scenario. We estimated that shifting a quarter of short (<5km) car trips to cycling would further prevent approximately 1,800 annual deaths and avoid €2.5 billion intangible costs. Table 4 further presents a broad comparison of elements to put into context the results of the modal shifts scenarios we assessed here. As a rough comparison, this is roughly the number of deaths avoided due to efforts in road safety made in France over the past 10 years, which represented a public investment of more than €3.5 billion a year ^46^. From a climate change perspective, shifting 25% of short car trips to cycling would yield a comparable CO_2_ emissions abatement than the 2015-2016 energy efficiency tax credit for households’ investment in home thermal renovation ^47^. However, it is also important to note that investments to foster active mobility may possibly result in large additional indirect cuts in CO_2_ emissions, for instance through encouraging multi-modal trips and/or re-location of activities which would reduce global distances travelled. A growing body of evidence has described how public policies relying on car use restriction, infrastructure development and incentives may effectively increase the share of active transportation ^48,49^.

**Table 4:** The public health, health-related economic and climate mitigation benefits of cycling in context, France.

| Dimension | Outcome | Value | Reference |
| --- | --- | --- | --- |
| Public health | Expected deaths yearly prevented by shifting 25% of short (<5km) car trips to bike | Approx.. 1,800 deaths yearly | Present study |
|  | Deaths prevented by recent efforts for road safety over the past 10 years | Approx.. 1,500 deaths yearly | <sup>46</sup> |
|  | Deaths prevented yearly by reducing alcohol consumption by 20% | Approx.. 1,500 deaths yearly | <sup>55</sup> |
| Economics | Expected medical (tangible) costs prevented by shifting 25% of short (<5km) car trips to bike | Approx. €100 million yearly | Present study |
|  | Intangible costs prevented by shifting 25% of short (<5km) car trips to bike | Approx. €2,600 million yearly | Present study |
|  | Yearly budget of the French National Cancer Institute (INCa) | €118.5 million in 2023 | https://www.e-cancer.fr/Institut-national-du-cancer/Qui-sommes-nous/Budget |
| Climate change mitigation | CO<sub>2</sub> emissions prevented by shifting 25% of short (<5km) car trips to bike (scenario 1) | Approx. 0.25 Mto yearly | Present study |
|  | Energy efficiency tax credit for households' investment in home thermal renovation | Approx. 0.12 Mto yearly in 2015 and 2016 | <sup>47</sup> |
|  | CO<sub>2</sub> emissions prevented by reducing the maximal speed on highway from 130 to 110 km/h | Approx. 1.45 Mto yearly | <sup>56</sup> |

Our study suffers from several limitations. First, we disregarded the levels of other types of physical activity when assessing the health benefits of existing or projected levels of cycling. However, the DRF we used came from meta-analyses of studies conducted in samples of participants with heterogeneous levels of physical activity and that controlled for various other domains of physical activity. Second, we did not account for the reduced physical activity that e-bikes may represent as compared to regular bikes. However, e-bikes only contributed to 6% of the distances we considered, suggesting a modest over-estimate of the benefits we document. Similarly, the modal shift scenarios we assessed did not account for a possible compensation during non-transport physical activity which could reduce the benefits we assessed. However, the existing evidence suggests that such substitution effect is unlikely or limited, in healthy adults at least ^50,51^. On the other hand, the dose-response relationship we used for all-cause mortality may be considered as conservative, as a more recent meta-analysis suggested a substantially more beneficial dose-response relationship ^52^. In a previous assessment, the choice of this dose-response relationship was identified as the main source of sensitivity in the estimates ^19^. Lastly, we did not explore the differential health impact of cycling across population groups, beyond gender. This limitation is common in similar studies ^12^, and at least partly due to the lack of reliable age-specific incidence and mortality data stratified by socio-economic position.

This study is one of the few to assess the benefits of cycling at the country level based on detailed individual transportation data and, to our knowledge, the first to do so for France. One of its main strengths lies in the fact that it documents both the medical and the social costs prevented by cycling. Although they disregard a major part of the estimated health benefits (those related to the mortality alleviated), medical costs represent tangible costs effectively saved for collective benefit. On the other hand, social costs based on VSLY are largely considered as intangible, because they represent the propensity of society to pay for the corresponding health benefits, but capture much more comprehensively the benefits expected from specific policies. The ratio of 1:25 we document here for these tangible and intangible costs may be useful to make sense of these figures in similar assessments. We hope that providing national estimates based on a representative survey and informed with reliable costs data, together with elements of contextualization as those presented in Table 4, could effectively inform national and local health and transportation policies.

The present study contributes to highlight the public health and climate mitigation benefits expected from the development of active transportation ^19,53^. Our results suggest that public investments to encourage modal shift toward cycling may translate into important climate, health and health-related economic benefits, which are likely to exceed the costs implied ^18^. Recently, due to the Covid-19 impact on public transportation, some local authorities have rapidly incentivized cycling by rolling out pop-up bike lanes. This resulted in a large short-term increase in cycling, including in France ^54^. Commitment of national and local authorities is critical to sustaining these changes and the contribution of cycling to public and planetary health.

## Data Availability

All data produced in the present study are available upon reasonable request to the authors.

https://www.statistiques.developpement-durable.gouv.fr/resultats-detailles-de-lenquete-mobilite-des-personnes-de-2019

## Contributors

ES, PQ, and KJ conceived the original idea. ML extracted and compiled input data. ES and KJ conducted the analysis and produced output figures and tables. ES, MS, AND, PQ, and KJ interpreted the results. ES and KJ wrote the first draft of the article. All authors provided critical feedback and helped shape the research, analysis and manuscript. All authors have approved the manuscript and gave their consent for submission and publication.

## Data sharing statement

Individual-level mobility data were accessed from the 2019 *Enquête mobilité des personnes* (“French People’s mobility survey”) at: https://www.statistiques.developpement-durable.gouv.fr/resultats-detailles-de-lenquete-mobilite-des-personnes-de-2019 [accessed April 5^th^ 2023.]. All data produced in the present study are available upon reasonable request to the authors.

## Ethics approval and consent to participate

Ethics approval and consent to participate have been collected by the institution in charge of the carrying out the survey: Ministère de la Transition écologique et de la Cohésion des territoires, Commissariat général au développement durable, Service des données et études statistiques (SDES) Sous-direction des statistiques des transports. The survey complies with all the standards of confidentiality of human data collection. More details here: https://www.statistiques.developpement-durable.gouv.fr/enquete-sur-la-mobilite-des-personnes-2018-2019?list-enquete=true

## Panel: Research in context

### Evidence before this study

We searched PubMed with the search equation (Bicycling[mesh] AND “health impact” and “physical activity”) on Nov 29^th^ 2023, with no date restriction. The research returned 35 articles. We also screened the reference lists of relevant articles returned by our search to identify other potentially relevant papers. A large part of the article assessed the health impact associated with scenarios projecting increasing cycling, alone or in combination with other features (eg increased walking or public transport use) and at varying scales (cities, metropolitan areas, countries). Other studies assessed the impact of specific schemes such as bike-sharing systems. Two studies only were conducted in France, one assessing the health benefits of reaching net-zero emission targets in transportation and the other simulating the impact of an urban mobility plan on physical activity, without reporting impacts on morbidity or mortality. We found only two studies reporting the impact of cycling both in terms of mortality, morbidity and costs saved: one for the UK (Woodcock et al) and the other in Melbourne, Australia (Brown et al). Overall, evidence shows that cycling may generate important population health and health economic benefits, though their magnitude largely varied across contexts and/or scenarios considered.

### Added value of this study

Based on pre-Covid nationally representative transportation data, we show that in a country of low- to moderate cycling culture such as France, cycling already generates important public health and health-related economic benefits, with an approximate annual 1,900 deaths and €5 billion of intangible costs averted. This corresponds to an approximate 1€ prevented for every km cycled. Currently, males are the main beneficiaries of cycling health benefits in France because they represent a large fraction of the cyclists.

Simulations based on individual transportation data shows that shifting only a quarter of short (<5km) car trips would yield approximatively a 2-fold increase in deaths prevented, while also contributing to more balanced health benefits across genders and generating sizeable CO_2_ emission reductions.

### Implication of all the available evidence

This study adds to the available evidence to show that cycling promotion policies would generate substantial public health benefits, which may be comparable to the gains expected by large-scale classical health prevention interventions, while also contributing to climate change mitigation targets.

