## Supplementary for "The untapped health and climate potential of cycling in France: a national assessment from individual travel data"

### Supplementary material 1 – Supplementary methods and results

**Supplementary table 1:** Relative risks relating physical activity and several chronic diseases as identified in a previous systematic review (Rojas-Rueda et al, 2013).

| Morbi-mortality event | Reference | RR (95% CI) | Unit | RR scaled for 100 min cycling, ie 11.25 MET.hours (95% CI) |
|---|---|---|---|---|
| Breast Cancer (women) | Monninkhof et al., 2007 | 0.94 (0.92, 0.97) | For each additional hour per week | 0.90 (0.87-0.95) |
| Colon Cancer | Harris et al., 2009 | Males: 0.80 (0.67, 0.96)<br><br>Females: 0.86 (0.76, 0.98) | Men: Per 30,1 METs per week<br><br>Women: Per 30,9 METs per week | Males: 0.93 (0.88-0.99)<br><br>Females: 0.94 (0.91-0.99) |
| Cardiovascular Disease | Hamer & Chida, 2008 | 0.84 (0.79,0.90) | 3 h per week of physical activity of moderate intensity | 0.92 (0.90-0.95) |
| Dementia | Hamer & Chida, 2009 | 0.72 (0.60, 0.86) | 33 METs per week | 0.90 (0.86-0.95) |
| Type 2 Diabetes | Jeon et al., 2007 | 0.83 (0.75, 0.91) | Per 10 METs per week | 0.81 (0.72-0.89) |

RR: relative risk; CI: confidence interval.
The scaled RR was obtained using the formula: $RR_{scaled}$ = 1-(1-$RR_{ref}$)*(11.25*$Unit_{ref}$).

**Supplementary table 2** : Estimates of disease mean duration and average discounted total medical costs

| **Disease** | **2018 number of prevalent cases (GBD 2019)** | **2018 number of incident cases (GBD 2019)** | **Mean duration (year)** | **Discounted mean duration* (year)** | **Average yearly medical costs (€$_{2018}$)** | **Average total medical costs (€$_{2018}$, accounting for discounting rate)** |
|---|---|---|---|---|---|---|
| Breast Cancer (females) | 581,096 | 50,787 | 11.44 | 9.95 | 4,720 | 46,968 |
| Colon Cancer | 278,713 | 50,936 | 5.47 | 5.11 | 5,224 | 26,716 |
| Cardiovascular Disease | 2,068,203 | 222,364 | 9.30 | 8.30 | 2,523 | 20,938 |
| Dementia | 1,104,927 | 150,914 | 7.32 | 6.69 | 3,400 | 22,748 |
| Type 2 Diabetes | 2,784,422 | 127,519 | 21.84 | 16.83 | 2,170 | 36,514 |

The method used to estimate average disease duration is those previously described in Dervaux et al (2022).

For each disease, the average duration by the ratio between the total number of prevalent cases and the total number of incident cases, provided by the 2019 Global Burden of Disease (https://vizhub.healthdata.org/gbd-results/). Such an estimation is made under the assumption of stationarity of the disease, ie assuming that disease prevalence and duration are stable over time. This estimated duration is then updated considering the discounting rate of 2.5%, which is the value recommended for the cost-benefit assessment of public investments in France (Quinet, 2013), and using the formula :
$D_{discounted} = (1 - \exp(-D * r)/D$; where *D* is the estimated undiscounted duration and *r* is the discounting rate.

Disease-specific average yearly medical costs were estimated based on expenses reimbursed by all health insurance schemes (available at: https://data.ameli.fr/explore/dataset/depenses/information/). These expenses, expressed in Euros 2018, include outpatient care, hospitalization in public or private healthcare facilities, and daily allowances.

Considering that the prevalence of chronic disease tends to increase and that medical progresses tend to increase disease duration, the assumption of disease stationarity may actually conduct to an under-estimation of disease duration, and therefore to an under-estimation of medical costs.

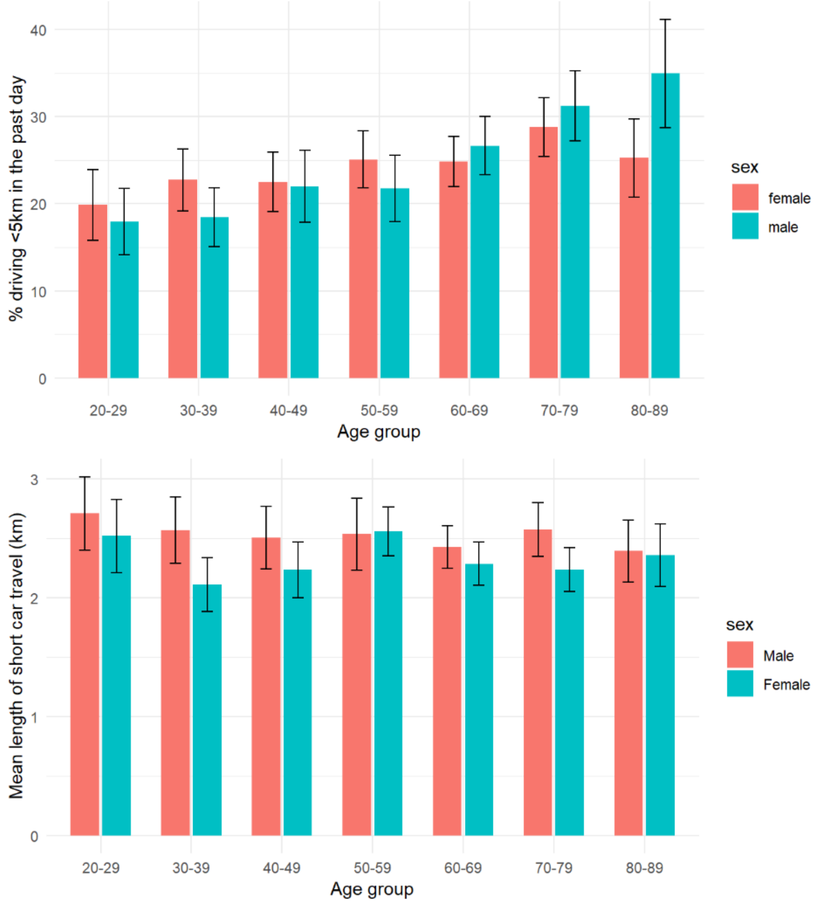


**Supplementary Figure 1:** Proportion of the French adult population reporting any short (<5km) car trip in the past day (top), and mean distance driven (km) in the past day among those reporting any car trip (down) according to sex and age. *Enquête mobilité des personnes,* France, 2019.
Black lines represent 95% confidence intervals.

**Supplementary table 3** : Climate, health and health-related economic benefits of cycling in France, 2019, and estimated impact of two modal shift scenarios.

| **Outcome** | **Baseline estimates (Uncertainty interval, UI)** | **Incremental effect of shifting 25% of short (<5km) car trips to cycling (in addition to the baseline estimates) (UI)** | **Incremental[1] effect of shifting 25% of short-to-medium (<10km) car trips to cycling (in addition to the baseline estimates) (UI)** |
|---|---|---|---|
| Yearly km cycled (billion) | 4.640 (3.284-5.996) | 2.073 (1.864-2.314) | 5.550 (4.222-6.884) |
| $CO_2$ emissions prevented (Mto) | 0.575 (0.4070.743) [1] | 0.257 (0.231-0.288) | 0.688 (0.524-0.854) |
| # of deaths prevented | 1,919 (1,101-2,736) | 1,822 (1,010-2,633) | 4,704 (2,689-6,721) |
| # of chronic diseases prevented | 5,963 (3,178-8,749) | 3,410 (2,343-4476) | 8,509 (5,205-11,813) |
| # DALYS prevented | 35,135 (22,693 – 48,791) | 19,493 (12,684-26,302) | 57,4650 (34,983-78,733) |
| Direct medical (tangible) costs savings (million €) | 191 (98-285) | 108 (71-144) | 267 (178-393) |
| Intangible costs prevented (billion €) | 4.75 (3.02-6.49) | 2.59 (1.69-3.50) | 7.56 (4.65-14.47) |
| Intangible costs prevented for every km cycled (€) | 1.01 (0.60-1.59) | 1.25 (0.82-1.73) | 1.37 (0.48-2.39) |

Intangible costs are estimated based on the value of a statistical life year (VSLY).

[1] As compared to a counterfactual where individual would have done the same trips driving instead of cycling
